# Minimal-Input Deep Learning for Remote Screening of REM Sleep Behavior Disorder

**DOI:** 10.1101/2025.07.01.25330646

**Authors:** Khrystyna Semkiv, Walter Karlen

## Abstract

This work investigates whether a deep learning model with minimal inputs can accurately identify Rapid Eye Movement Sleep Behavioral Disorder (RBD). We propose an interpretable two-step approach using two convolutional neural networks for sleep staging and RBD classification. Experiments on data from 18 RBD participants and 178 healthy controls demonstrate that reliable classification can be achieved using frontal electroencephalogram (EEG) and electrooculogram (EOG) input signals. GradCAM attention reveals a 22% increase in importance in the 9-22 Hz band of EOG for RBD cases. Our findings highlight the potential for remote, wearable-based RBD screening at home.

## 1 Introduction

Rapid eye movement (REM) sleep behavior disorder (RBD) is a parasomnia characterized by a lack of muscle paralysis during REM sleep, leading to dream-related motor activity [1]. Isolated, formerly known as idiopathic, RBD is considered an early phase synucleinopathy because within 12 years after diagnosis, up to 73.5% of cases will progress to neurodegenerative synucleinopathy, such as Parkinson’s disease or Lewy body dementia [2]. An accurate and timely diagnosis of isolated RBD can provide an early entry point for the screening of neurodegenerative disorders.

A well-established approach for RBD diagnosis is overnight video-polysomnography (video-PSG). In the sleep lab, REM sleep is classified from electrophysiological recordings and then, these REM episodes are screened for abnormal behavior in the electromyogram (EMG) and video [3]. However, this approach is costly and not easily accessible, as it can only be provided by trained staff in specialized centers who perform time-consuming manual analysis, often including subjective assessments [4]. Therefore, a more appropriate, scalable approach for RBD screening, which is accessible, accurate, objective, and automated, is needed.

Automation of RBD screening from PSG recordings with machine learning (ML) to reduce complexity and improve objectivity has been already demonstrated. For example, Brink-Kjaer et al. introduced an end-to-end deep learning model that uses solely two electroencephalogram (EEG) channels, achieving an overall accuracy of 84.5% [5]. In a two-step approach, consisting of sleep staging and RBD classification, Cooray et al. explored the PSG channels of EEG, EMG, and electrooculography (EOG) with a Random Forest (RF) classifier, achieving 92% accuracy [6]. In their next work, they replaced EEG with an electrocardiography signal and explored various channel configurations, achieving 90% using EOG and EMG [7]. However, these models focused on input channels that originate from PSG recordings only. Additionally, EEG channels originated from central derivations (C4-A1, C3-A2) [5, 6] which are difficult to obtain from a self-applied wearable due to the abundance of hair and restricted visibility in the mirror of these locations. Furthermore, the limb EMG channel used in [5] would require extended cabling or a multi-device setup. Considering these limitations, more research is needed to design a patient-friendly device that requires a minimal number of recording channels and offers comfortable placement to minimize disruption of natural sleep at home.

In this study, we investigate whether a fully automated approach using deep learning can detect the presence of RBD from signals available from a wearable (Figure 1) and whether the model can be reduced to one channel to simplify the setup. Our direct contributions to the field of RBD screening are: 1) Development of an automated two-step deep learning approach mimicking clinical procedures and enabling interpretation of results; 2) Development of a convolutional neural network (CNN) with spectrogram input, attention mechanisms, and threshold-based fine-tuning; and 3) Identification of the minimal number of recording channels needed on a wearable to perform screening outside of the sleep lab.

**Figure 1.**
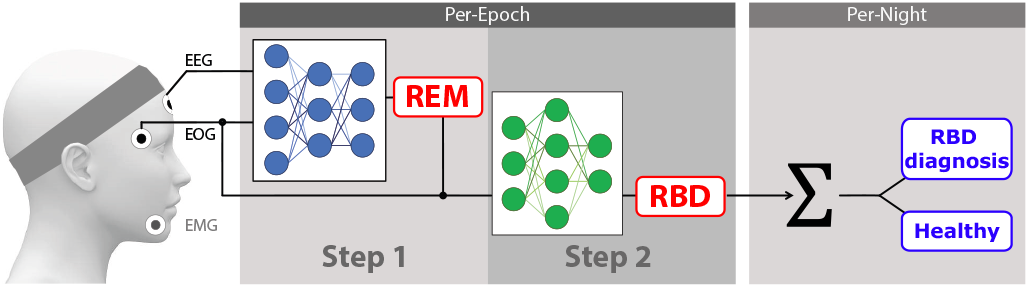
Fully automated RBD screening with two-step deep learning from a minimal set of electrophysiological signals obtained with a wearable.

## 2 Materials and Methods

### 2.1 Design of models and pipeline

Inspired by clinical practice, we developed a fully automated deep learning approach consisting of two distinct steps that provide more transparency and modularity over a single, large model. In the first step, we integrated sleep staging using an established open-source model, with the main purpose to localize REM sleep epochs. For the second step, we designed a CNN model to analyze the time-frequency characteristics of the electrophysiological signals within the REM sleep epochs and identify RBD specific attributions.

For the sleep staging step, we selected the U-Sleep model, which was designed for cross-dataset generalization and a multitude of health conditions [8]. We chose 30-second epochs of EEG and EOG as input for the model, which predicted 4 sleep and wake stages to create a hypnogram.

For the RBD classification step, we custom-built a CNN-based feature extraction model with spectrograms as input. Representation learning started with an initial large kernel size convolutional layer to learn global features across the input, followed by a strided convolutional layer for dimensionality reduction (Figure 2 a). Four inverted residual (IR) bottleneck blocks [9] were attached to learn more complex and local features (Figure 2 b). The computationally lightweight IR blocks were composed of three layers: a convolution layer to temporarily expand feature space, a depthwise convolution layer to operate in expanded feature space, and one more convolution layer to compress the information back down. The output of the last IR block was passed through a fully connected (FC) layer to generate per-epoch predictions. Additionally, after the last IR block, we integrated a localized GradCAM mechanism which estimated the importance values for each input spectrogram [10].

**Figure 2.**
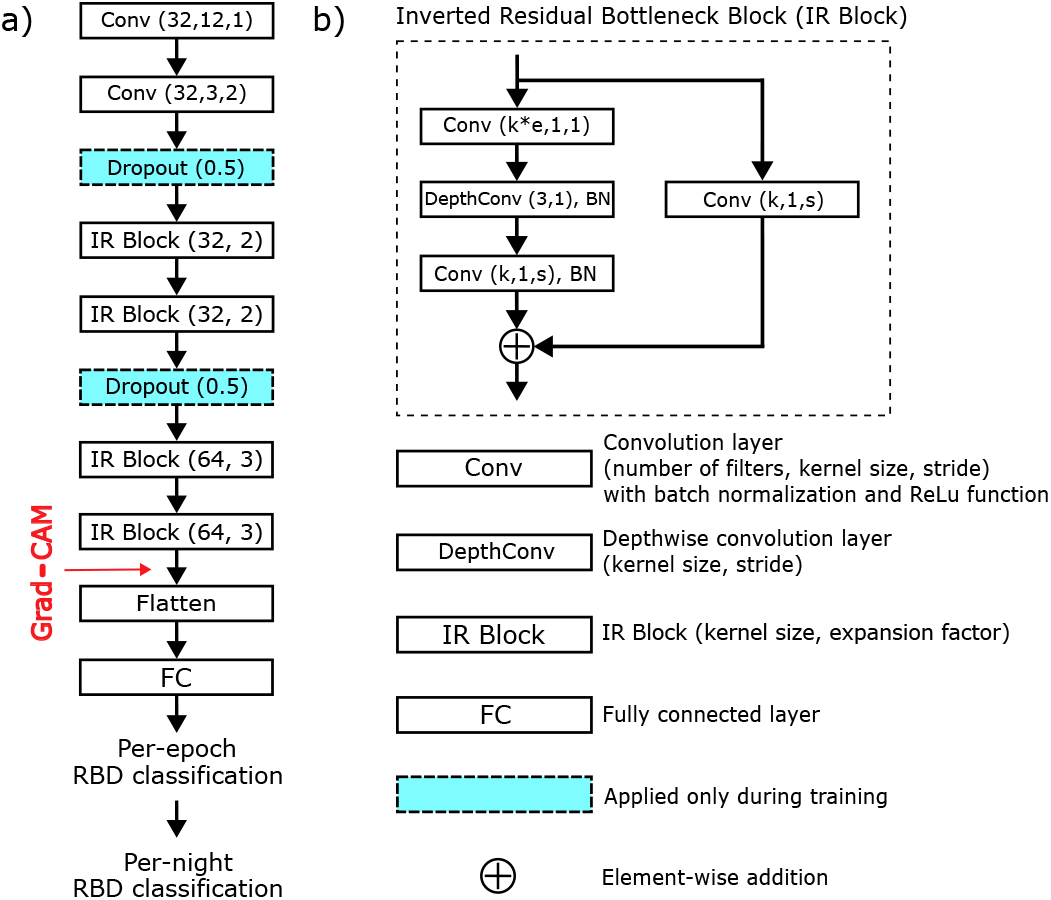
The CNN-based model for RBD classification: a) full architecture, and b) IR block. *BN* denotes batch normalization. *e* denotes an expansion factor of the feature space in the IR block.

To receive per-night predictions, we accumulated the duration of all RBD classified epochs. A learned decision threshold then classified recordings for RBD diagnosis.

### 2.2 Datasets and data preparation

For the experiments, we aggregated data from three different sources: crease the robustness of the models against acquisition artifacts, we the Cyclical Alternating Pattern (CAP) sleep database [11, 12], the WESA ambulatory sleep recordings that used the MHSL-SleepBand [13], and the Montreal Archive of Sleep Studies (MASS) [14]. From the recordings, we extracted the channels suitable for wearable devices: frontal EEG (Fpz or closest), Δ*EOG*_*right−left*_, and chin submentalis EMG. We excluded recordings that were duplicates, unreadable, too noisy, had missing channels, or originated from participants with severe sleep apnea (AHI ¿ 30). Finally, we age-matched (am) the remaining 178 healthy control (HC) recordings to the RBD group: RBD_*CAP*_ (18), 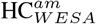 (12), 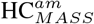(53), HC_*CAP*_ (6), HC_*MASS*_ (107).

To ensure homogeneous data across sources, each signal was passed through a Butterworth filter, with order and cut-off frequencies recommended by the AASM Manual [15]: EEG (4^*th*^-order, 0.1-40 Hz), EMG (5^*th*^-order, 10-100 Hz), and EOG (4^*th*^-order, 0.1-35 Hz). Power line noise was removed with a notch filter and all signals were resampled to 250 Hz. The manual labels were resampled to 30-second epochs.

The recordings were split into training (107 HC_*MASS*_ and 9 RBD_*CAP*_), validation (30 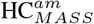 and 4 RBD_*CAP*_), adaptation (6 HC_*CAP*_), and test (23 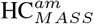, 12 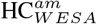, and 5 RBD_*CAP*_) sets. We isolated REM epochs of the CAP using expert labels and applied a synthetic minority oversampling technique (SMOTE) [16] to augment the sets with additional synthetic RBD recordings (RBD_*SY N*_) in training (100), validation (26), and adaptation (30) as well as 24 HC_*SY N*_ in adaptation. Finally, we transformed all epochs into spectrograms using short-time Fourier transformation with a 2-second Hann window and 90% overlap.

### 2.3 Experiments

We conducted ML experiments following the ethical principles of the Declaration of Helsinki. Data reuse and experiments were approved by the local ethics board (ID 231/24). We ran two experiments to determine the minimal number of channels needed for RBD screening, and evaluate the performance of our fully automated, two-step approach.

#### 2.3.1 Input channel minimization

This experiment aimed to identify the minimal number of recording channels for a reliable RBD classification. For this, we trained and validated 6 models with variable input configurations. Crossentropy loss was minimized using an Adam optimizer over 100 epochs. The batch size was set to 128 with the learning rate 0.5 × 10^*−*5^. The models were equipped with regularization mechanisms such as L1, L2, and a dropout. Furthermore, we applied a gradient clipping and an early stopping after 10 consecutive epochs. To increase the robustness of the models against acquisition artifacts, we re-trained it with the adaptation set using 6-fold cross-validation.

To determine an optimal decision threshold for the per-night RBD diagnosis, we performed a Receiver Operating Curve optimization on the re-trained model with the best generalization performance. The threshold was tuned from 30 s to the maximum possible REM duration overnight with an increment of 0.5 s. The geometric mean between specificity, the proportion of correctly predicted HC cases, and sensitivity, the proportion of correctly predicted RBD cases, was maximized. Recordings with total REM duration shorter than the decision threshold were excluded from per-night evaluation.

#### 2.3.2 Fully automated RBD screening

This experiment aimed to assess the performance of the fully automated RBD screening pipeline (Figure 1) with the most promising input configuration from the previous experiment and identify the most salient features of the available signals.

#### 2.3.3 Evaluation

We estimated for each model and experiment specificity and sensitivity for per-epoch (mean *±* SD), as well as per-night predictions. We also reported the unified attention maps by averaging all REM attention maps produced by the GradCAM in the fully automated approach for the HC and RBD groups and visualized the difference to highlight possible features discriminating the group.

## 3 Results

### 3.1 Input channel minimization

The EOG channel achieved the highest per-epoch mean specificity at 94.9% with the lowest SD (Table 1). The EOG channel also excelled with a per-night specificity of 97.7% and sensitivity of 100% (N=45, 5 RBD).

**Table 1.** Per-epoch and per-night specificity (Sp) and sensitivity (Se) of RBD classification with expert and automated sleep staging.

| Model<br>Input | Per-epoch |  | Per-night |  |
| --- | --- | --- | --- | --- |
|  | Sp (%) | Se (%) | Sp (%) | Se (%) |
| **Experiment 1: Expert sleep staging** |  |  |  |  |
| EEG | 31.8 ± 45.7 | 66.6 ± 50.1 | 0.0 | 100.0 |
| EMG | 70.2 ± 13.9 | 55.7 ± 13.2 | 84.4 | 80.0 |
| EOG | 94.9 ± 2.2 | 63.9 ± 17.3 | 97.7 | 100.0 |
| EEG EMG | 78.2 ± 14.1 | 40.8 ± 24.7 | 82.2 | 100.0 |
| EEG EOG | 69.5 ± 18.2 | 48.2 ± 30.7 | 62.2 | 100.0 |
| EMG EOG | 83.1 ± 6.2 | 69.0 ± 21.9 | 97.7 | 80.0 |
| **Experiment 2: Automated sleep staging** |  |  |  |  |
| EOG | 95.7 | 65.3 | 100.0 | 100.0 |

#### 3.2 Fully automated RBD screening

When assessing the classification of the whole diagnostic pipeline with EOG, no performance decline could be observed as per-epoch specificity (95.7%) and sensitivity (65.3%) were within the range of experiment 1 (Table 1). All recordings were correctly classified (N=44, 5 RBD). Furthermore, the attention maps showed a 22% increase in importance in the 9-22 Hz region for the RBD group (Figure 3).

**Figure 3.**
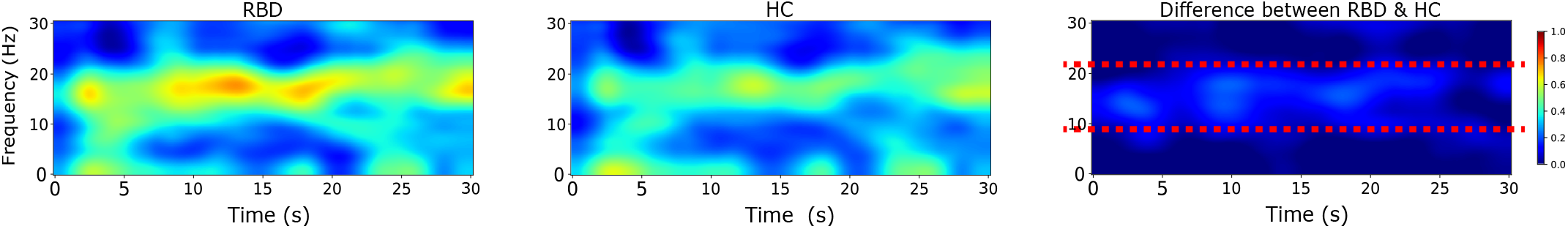
Attention maps for EOG spectrograms generated by the GradCAM mechanism on REM sleep epochs from RBD participants (RBD, left) and healthy controls (HC, middle) in the test set. Red dashed lines depict the frequency range with major differences between attention (right).

## 4 Discussion

In this work, we introduced a fully automated deep learning RBD screening and investigated whether a minimal setup can be found for wearable headwear. We found that two successive convolutional networks, REM sleep detection with EEG and EOG input followed by RBD classification with EOG input, can provide high sensitivity and specificity on a single screening night. Both input signals can be easily obtained from electrode derivations available in wearables for use at home.

We identified the single EOG input as the best performing channel for the per-epoch and per-night classification of RBD. Combined with the EEG channel which is needed for REM sleep detection in the first step, this is the minimal channel number we could identify that provides acceptable performance. Relying solely on EEG was not sufficient as it lead to high performance variation with a class overfitting. This is in contrast to Brink-Kjaer et al. who achieved a specificity of 82.0% and a sensitivity of 86.8% using a single central EEG channel with an end-to-end deep learning model [5]. As different EEG channel placement (frontal vs central), epoch length (30 s vs 300 s), and data sets were used, we cannot provide a conclusive statement regarding the suitability of using a single channel for RBD screening. A major limiting factor might have been the availability of only per-night labels of RBD in our set. This absence of behavioral labels for each 30-second epoch resulted in the labeling of all REM epochs, including those expressing normal behavior, with the RBD class. This over-labeling led to difficulties in the interpretation of the per-epoch performance and might have also weakened the representation learning, especially from the EEG signal. A solution to this challenge could be the inclusion of an evidence aggregation model that could perform temporal ensembling [17].

We tested the two-step RBD classification with the EOG channel input using automated sleep stages generated by U-Sleep. Per-epoch and per-night prediction performances did not drop (Table 1), demonstrating the effectiveness of a fully automated approach. The attention maps showed EOG activity around 9-22 Hz with higher intensity for RBD participants (Figure 3) which could indicate facial muscle atonia loss. Christensen et al. used wavelet decomposition to analyze EOG signals, and found distinct energy values in 16-64 Hz that distinguished idiopathic RBD from HC [18]. A sensor fusion approach, i.e. with additional movement or sound sensors, could provide further insights into whether the higher frequency activity reflects the loss of facial muscle atonia.

The main limitation of this retrospective study was the requirement to use existing data sets containing only a restricted number of RBD recordings. Notably, we had no RBD recordings obtained with the wearable head band. Therefore, we can not entirely exclude selection and data biases. As different recoding setups have been used for generating the data sets, we can’t exclude that the ML training also picked up characteristics reflecting differences in recording systems and setups. We tried to control for this with the re-training on a single data source. An additional limitation was the absence of information on RBD severity and phenotype which restricted the analysis to the per-night diagnostic performance, preventing a granular analysis with sub-group specifics.

In conclusion, fully automated two-step RBD screening is feasible with only two channels, EEG and EOG, which can easily be recorded with wearable devices. Further research with larger, prospectively collected datasets are needed.

## Data Availability

The data used in this study were obtained from three sources: (1) an open-access dataset publicly available with registration or usage terms; (2) a dataset available upon request from the original investigators; and (3) a private dataset not publicly available due to institutional or ethical restrictions. Access to the open and request-based datasets can be arranged through the sources cited in this manuscript.

